# Sex-Mismatching in Isolated Heart Transplant Confers No Postoperative Risk

**DOI:** 10.1101/2024.02.23.24303301

**Authors:** Reid Dale, Matt Leipzig, Nataliya Bahatyrevich, Katharine Pines, Qiudong Chen, Jeffrey Teuteberg, Y. Joseph Woo, Maria Currie

## Abstract

**Introduction:** For heart transplantation, optimal donor-recipient matching is an important factor in the ongoing development of the United Network for Organ Sharing (UNOS) continuous distribution framework. Donor-recipient sex-mismatch has decreased since the 1990s, but this may be related to the risk posed by size mismatching, particularly when donor hearts are undersized. Thus, the impact of sex-mismatching, controlling for other factors including size mismatch, is uncertain.

**Methods:** Adult first-time, isolated heart transplant patients from the UNOS database between October 1, 1987 and December 31, 2022 were analyzed. Cohorts were separated into male and female recipients. Propensity score matching on known preoperative risk factors was performed. Equivalence testing via Two One-Sided Testing (TOST) was performed to assess between-arm equivalence in postoperative outcomes. Survival differences were measured by the between-arm ratio of Restricted Mean Survival Time and binary outcome differences by the Odds Ratio (OR).

**Results:** In the propensity matched cohort, we found significant equivalence between arms in both male (TOST P<0.001) and female (TOST P<0.001) recipients for overall survival at all temporal endpoints, postoperative treatment for rejection within one year, and pre-discharge dialysis.

**Conclusions:** Sex-mismatch in isolated heart transplantation confers no additional risk to postoperative outcomes when controlling for other factors, including size mismatch. Consequently, sex-mismatch should not factor into individual assessments of organ acceptance or be incorporated into any national organ allocation policy. Increasing the acceptance of sex-mismatched donors has the potential to expand the donor pool and increase female donor utilization.

## Introduction

Heart transplantation is an established treatment for end-stage heart failure. There are over 5.8 million patients with heart failure in the USA and this number is expected to continue to increase.^1^ Of those with heart failure, over 10% have end-stage disease.^1,2^ Even though the total number of available donor hearts has steadily increased since 2011, the acceptance rate of donor hearts has decreased to 30% in 2018 from its peak of 45% in 1995.^3^

The risk profile of donor-recipient sex-mismatching has been a focal point of recent research including all-cause mortality, stroke, dialysis, and graft dysfunction. There have been conflicting findings regarding the risk that sex-mismatching poses as a function of recipient sex.^4^ For example, a 2012 retrospective study observed a significant negative association between sex-mismatch and survival for both male and female recipients.^5^ However, a later 2014 study drawing from the United Network for Organ Sharing (UNOS) Transplant Database argued that the observed negative association between sex-mismatch and survival for male recipients was instead due to donor-recipient heart size mismatch.^6^ Complicating matters further, a 2017 study found that after controlling for height and weight mismatch between donor and recipient, sex-mismatching posed a risk to both male and female recipients.^7^

Given the conflicting findings within the literature, re-evaluation of sex-mismatch risk using a novel statistical design is warranted. We aim to definitively resolve the question of the clinical relevance of sex-mismatching on a recipient’s postoperative outcomes through the use of a design combining methods from equivalence testing and causal inference.

## Methods

### Patient Population

We selected patients undergoing heart transplantation in the UNOS database from October 1^st^ 1987 to December 31^st^ 2022. We excluded recipients under 18 at the time of waitlisting, multiorgan transplant, and retransplants. We then separated the cohort between male and female recipients to replicate the findings of previous studies and to control for base rate differences between recipient sexes in heart size and differences in the availability of organs.

Recipient characteristics, donor characteristics, and patient outcomes were defined according to the standard UNOS definitions. Donor to recipient Predicted Heart Mass (PHM) ratio was calculated with a previously developed formula using recipient age, sex, height, and weight.^8^ PHM is a commonly used surrogate for donor-recipient size match.

### Primary and Secondary Outcomes

The primary outcome was overall postoperative survival. The secondary outcomes included stroke prior to discharge, dialysis prior to discharge, and treatment for acute rejection within one year.

### Statistical Analysis

#### Descriptive Reporting and Statistical Estimation Procedures

We reported baseline covariates in the unmatched cohorts for each recipient sex stratified by sex-mismatch (Table 1). Propensity score matching was used to create balanced groups of sex-matched and sex-mismatched recipients. We selected imbalanced variables that would affect the propensity of sex-mismatching to generate a propensity score for each patient. These variables were: recipient age at time of transplant, recipient body mass index (BMI), transplant year, donor-recipient PHM ratio, UNOS priority status at time of transplant, etiology of heart failure, ventricular assist device utilization and type at time of transplant, extracorporeal membrane oxygenation at time of transplant, intra-aortic balloon pump at time of transplant, life support at time of transplant, ischemic time, donor age, and donor ejection fraction. We performed nearest neighbor matching with a standardized mean difference (SMD) caliper of 0.2 using a random forest model of propensity. Matched variables with an SMD at or below 0.25 and variance ratios between 0.5 and 2.0 were considered well-matched in accordance with the benchmarks outlined by Stuart.^9^ We plotted the probability of the donor-recipient PHM ratio for sex-matched and sex-mismatched patients stratified by recipient sex. Values greater than one indicate that the donor had a larger PHM than the recipient, one indicates no difference, and less than one means that the recipient PHM was larger than the donor. In addition, we tested the trends in utilization of sex-mismatching versus sex-matching across years stratified by recipient sex via the Cochrane-Armitage trend test.

**Table 1:** Recipient Demographics: All-Comers.

| Variable | Male Recipients |  |  | Female Recipients |  |  |
| --- | --- | --- | --- | --- | --- | --- |
|  | Sex-Matched,<br>N = 42,446<sup>1</sup> | Sex-Mismatched,<br>N = 11,149<sup>1</sup> | Difference<sup>2</sup> | Sex-Matched,<br>N = 9,443<sup>1</sup> | Sex-Mismatched,<br>N = 7,572<sup>1</sup> | Difference<sup>2</sup> |
| **Donor/Recipient PHM Ratio** | 1.01 (0.93, 1.10) | 0.83 (0.76, 0.91) | 1.4 | 1.03 (0.95, 1.14) | 1.29 (1.17, 1.42) | -1.4 |
| Unknown | 4,411 | 1,417 |  | 685 | 758 |  |
| **Recipient Age, y** | 55 (47, 62) | 56 (48, 62) | -0.05 | 53 (42, 60) | 52 (40, 59) | 0.12 |
| **Recipient Race** |  |  | 0.17 |  |  | 0.06 |
| Asian, Non-Hispanic | 910 (2.1%) | 451 (4.0%) |  | 247 (2.6%) | 147 (1.9%) |  |
| Black, Non-Hispanic | 6,654 (16%) | 1,298 (12%) |  | 2,186 (23%) | 1,902 (25%) |  |
| Hispanic/Latino | 2,821 (6.6%) | 927 (8.3%) |  | 711 (7.5%) | 537 (7.1%) |  |
| Other | 349 (0.8%) | 78 (0.7%) |  | 86 (0.9%) | 76 (1.0%) |  |
| White, Non-Hispanic | 31,696 (75%) | 8,389 (75%) |  | 6,209 (66%) | 4,908 (65%) |  |
| Unknown | 16 | 6 |  | 4 | 2 |  |
| **Diabetes mellitus** | 9,169 (26%) | 2,060 (24%) | 0.06 | 1,715 (21%) | 1,273 (20%) | 0.01 |
| Unknown | 7,766 | 2,452 |  | 1,149 | 1,318 |  |
| **Total waitlist time, days** | 93 (26, 266) | 72 (21, 210) | 0.12 | 61 (18, 186) | 63 (17, 192) | 0.00 |
| **Prior cardiac surgery, n (%)** | 12,607 (30%) | 2,729 (24%) | 0.12 | 2,536 (27%) | 1,700 (22%) | 0.10 |
| **Prior lung surgery, n (%)** | 316 (0.7%) | 117 (1.0%) | 0.21 | 49 (0.5%) | 51 (0.7%) | 0.20 |
| **Hospitalization status at time of transplant** |  |  | 0.02 |  |  | 0.20 |
| HOSPITALIZED NOT IN ICU | 6,158 (15%) | 1,544 (14%) |  | 1,247 (13%) | 1,144 (15%) |  |
| IN INTENSIVE CARE UNIT | 16,920 (40%) | 4,468 (40%) |  | 3,204 (34%) | 3,181 (42%) |  |
| NOT HOSPITALIZED | 19,012 (45%) | 5,067 (46%) |  | 4,894 (52%) | 3,206 (43%) |  |
| Unknown | 356 | 70 |  | 98 | 41 |  |
| **Blood type, n (%)** |  |  | 0.05 |  |  | 0.07 |
| A | 17,910 (42%) | 4,870 (44%) |  | 3,841 (41%) | 2,952 (39%) |  |
| AB | 2,347 (5.5%) | 512 (4.6%) |  | 409 (4.3%) | 428 (5.7%) |  |
| B | 5,980 (14%) | 1,509 (14%) |  | 1,303 (14%) | 1,128 (15%) |  |
| O | 16,165 (38%) | 4,250 (38%) |  | 3,880 (41%) | 3,062 (40%) |  |
| Unknown | 44 | 8 |  | 10 | 2 |  |
| **Total bilirubin, mg/dL** | 1.15 (2.26) | 1.22 (2.57) | -0.03 | 1.03 (2.55) | 1.08 (2.18) | -0.02 |
| Unknown | 8,072 | 2,599 |  | 1,339 | 1,354 |  |
| **Transplant Year** | 2,007 (1,997, 2,016) | 2,003 (1,995, 2,014) | 0.22 | 2,011 (2,000, 2,018) | 2,006 (1,997, 2,015) | 0.26 |
| **Weight (kg)** | 85 (75, 97) | 76 (68, 86) | 0.57 | 66 (57, 78) | 69 (59, 81) | -0.21 |
| Unknown | 234 | 83 |  | 54 | 46 |  |
| **Height (cm)** | 178 (173, 183) | 175 (170, 180) | 0.51 | 163 (157, 168) | 164 (160, 168) | -0.25 |
| Unknown | 299 | 100 |  | 55 | 58 |  |
| **Waitlist priority status at time of transplant** |  |  | 0.30 |  |  | 0.34 |
| HR: Adult Status 1 | 803 (1.9%) | 56 (0.5%) |  | 89 (1.0%) | 150 (2.0%) |  |
| HR: Adult Status 2 | 3,947 (9.5%) | 505 (4.6%) |  | 736 (7.9%) | 590 (8.0%) |  |
| HR: Adult Status 3 | 1,258 (3.0%) | 253 (2.3%) |  | 387 (4.2%) | 172 (2.3%) |  |
| HR: Adult Status 4 | 1,107 (2.7%) | 343 (3.2%) |  | 622 (6.7%) | 202 (2.7%) |  |
| HR: Adult Status 5 | 2 (<0.1%) | 1 (<0.1%) |  | 3 (<0.1%) | 0 (0%) |  |
| HR: Adult Status 6 | 293 (0.7%) | 87 (0.8%) |  | 212 (2.3%) | 50 (0.7%) |  |
| HR: Old Status 1 | 6,615 (16%) | 2,217 (20%) |  | 936 (10%) | 1,208 (16%) |  |
| HR: Status 1A | 12,489 (30%) | 2,772 (25%) |  | 2,263 (24%) | 2,064 (28%) |  |
| HR: Status 1B | 7,794 (19%) | 2,146 (20%) |  | 2,104 (23%) | 1,523 (21%) |  |
| HR: Status 2 | 6,552 (16%) | 2,343 (22%) |  | 1,884 (20%) | 1,372 (18%) |  |
| HR: Temporarily inactive | 577 (1.4%) | 161 (1.5%) |  | 69 (0.7%) | 89 (1.2%) |  |
| Unknown | 1,009 | 265 |  | 138 | 152 |  |
| **VAD Device Type** |  |  | 0.18 |  |  | 0.16 |
| LVAD | 9,845 (36%) | 1,744 (28%) |  | 1,665 (25%) | 1,157 (24%) |  |
| LVAD/RVAD/TAH Unspecified | 1,991 (7.2%) | 505 (8.1%) |  | 228 (3.4%) | 298 (6.2%) |  |
| LVAD+RVAD | 627 (2.3%) | 108 (1.7%) |  | 115 (1.7%) | 127 (2.7%) |  |
| NONE | 14,866 (54%) | 3,821 (61%) |  | 4,644 (69%) | 3,147 (66%) |  |
|  | Sex-Matched,<br>N = 42,446<sup>1</sup> | Sex-Mismatched,<br>N = 11,149<sup>1</sup> | Difference<sup>2</sup> | Sex-Matched,<br>N = 9,443<sup>1</sup> | Sex-Mismatched,<br>N = 7,572<sup>1</sup> | Difference<sup>2</sup> |
| RVAD | 68 (0.2%) | 16 (0.3%) |  | 12 (0.2%) | 13 (0.3%) |  |
| TAH | 246 (0.9%) | 30 (0.5%) |  | 19 (0.3%) | 31 (0.6%) |  |
| Unknown | 14,803 | 4,925 |  | 2,760 | 2,799 |  |
| **ECMO** | 548 (1.3%) | 84 (0.8%) | 0.05 | 98 (1.0%) | 114 (1.5%) | -0.04 |
| **IABP** | 4,160 (9.8%) | 942 (8.4%) | 0.05 | 855 (9.1%) | 718 (9.5%) | -0.01 |
| **Etiology of Heart Failure** |  |  | 0.09 |  |  | 0.05 |
| Congenital | 831 (2.0%) | 277 (2.5%) |  | 391 (4.2%) | 293 (3.9%) |  |
| exclude | 85 (0.2%) | 23 (0.2%) |  | 20 (0.2%) | 15 (0.2%) |  |
| Idiopathic | 14,581 (34%) | 3,440 (31%) |  | 3,788 (40%) | 3,209 (42%) |  |
| Ischemic | 20,008 (47%) | 5,642 (51%) |  | 2,039 (22%) | 1,652 (22%) |  |
| Other | 6,877 (16%) | 1,752 (16%) |  | 3,178 (34%) | 2,390 (32%) |  |
| Unknown | 64 | 15 |  | 27 | 13 |  |
| **Cockcroft Creatinine Clearance** | 817 (632, 1,053) | 737 (566, 952) | 0.22 | 693 (529, 916) | 723 (544, 976) | -0.11 |
| Unknown | 6,811 | 2,194 |  | 1,109 | 1,133 |  |
| **UNOS Region** |  |  | 0.15 |  |  | 0.12 |
| 1 | 1,771 (4.2%) | 531 (4.8%) |  | 426 (4.5%) | 337 (4.5%) |  |
| 2 | 4,968 (12%) | 1,528 (14%) |  | 1,205 (13%) | 783 (10%) |  |
| 3 | 5,281 (12%) | 1,099 (9.9%) |  | 1,119 (12%) | 948 (13%) |  |
| 4 | 4,480 (11%) | 1,142 (10%) |  | 907 (9.6%) | 820 (11%) |  |
| 5 | 6,731 (16%) | 1,636 (15%) |  | 1,281 (14%) | 1,214 (16%) |  |
| 6 | 1,333 (3.1%) | 296 (2.7%) |  | 319 (3.4%) | 238 (3.1%) |  |
| 7 | 3,813 (9.0%) | 1,056 (9.5%) |  | 879 (9.3%) | 663 (8.8%) |  |
| 8 | 2,727 (6.4%) | 662 (5.9%) |  | 594 (6.3%) | 480 (6.3%) |  |
| 9 | 2,482 (5.8%) | 911 (8.2%) |  | 639 (6.8%) | 420 (5.5%) |  |
| 10 | 3,671 (8.6%) | 1,080 (9.7%) |  | 902 (9.6%) | 694 (9.2%) |  |
| 11 | 5,189 (12%) | 1,208 (11%) |  | 1,172 (12%) | 975 (13%) |  |
| **Donor Age** | 28 (21, 38) | 35 (25, 45) | -0.41 | 33 (23, 43) | 25 (19, 36) | 0.48 |
| Unknown | 1 | 1 |  |  |  |  |
| **Donor Sex** |  |  | 0.00 |  |  | 0.00 |
| Female | 0 (0%) | 11,149 (100%) |  | 9,443 (100%) | 0 (0%) |  |
| Male | 42,446 (100%) | 0 (0%) |  | 0 (0%) | 7,572 (100%) |  |
| **Donor Height (cm)** | 178 (174, 183) | 168 (163, 170) | 1.7 | 163 (158, 168) | 175 (169, 180) | -1.3 |
| Unknown | 4,220 | 1,348 |  | 638 | 722 |  |
| **Donor Weight (kg)** | 82 (73, 94) | 74 (64, 89) | 0.35 | 67 (58, 80) | 72 (64, 82) | -0.19 |
| Unknown | 2,010 | 550 |  | 284 | 312 |  |
| **Calculated Donor BMI** | 25.7 (23.0, 29.2) | 26.9 (23.3, 32.1) | -0.29 | 25.3 (22.0, 30.0) | 23.7 (21.2, 26.8) | 0.40 |
| Unknown | 4,235 | 1,352 |  | 648 | 728 |  |
| **Donor LV Ejection Fraction** | 60 (55, 65) | 60 (57, 65) | -0.08 | 60 (57, 65) | 60 (55, 65) | 0.07 |
| Unknown | 13,365 | 4,455 |  | 2,270 | 2,468 |  |
| **Ischemic Time (hours)** | 3.05 (2.33, 3.73) | 3.10 (2.37, 3.77) | -0.02 | 3.17 (2.45, 3.82) | 3.00 (2.27, 3.67) | 0.13 |
| Unknown | 1,619 | 459 |  | 346 | 327 |  |
| **Diabetes mellitus** | 908 (2.6%) | 373 (4.2%) | -0.09 | 323 (3.9%) | 137 (2.2%) | 0.10 |
| Unknown | 7,042 | 2,188 |  | 1,060 | 1,200 |  |
| **Donor Hypertension** | 4,224 (12%) | 1,830 (20%) | -0.23 | 1,532 (18%) | 534 (8.4%) | 0.29 |
| Unknown | 7,099 | 2,211 |  | 1,068 | 1,217 |  |
| **Donor total bilirubin, mg/dL** | 0.80 (0.50, 1.30) | 0.60 (0.40, 1.00) | 0.12 | 0.60 (0.40, 1.00) | 0.80 (0.50, 1.30) | -0.11 |
| Unknown | 7,253 | 2,282 |  | 1,120 | 1,239 |  |
| **Blood type donor, n (%)** |  |  | 0.04 |  |  | 0.08 |
| A | 15,806 (37%) | 4,239 (38%) |  | 3,280 (35%) | 2,563 (34%) |  |
| AB | 1,069 (2.5%) | 215 (1.9%) |  | 130 (1.4%) | 178 (2.4%) |  |
| B | 4,584 (11%) | 1,172 (11%) |  | 911 (9.6%) | 820 (11%) |  |
| O | 20,986 (49%) | 5,522 (50%) |  | 5,121 (54%) | 4,010 (53%) |  |
| Unknown | 1 | 1 |  | 1 | 1 |  |

<sup>1</sup>Median (IQR); n (%); Mean (SD)
<sup>2</sup>Standardized Mean Difference

To gauge regional differences in sex-mismatching for both male and female recipients in the contemporary era beginning with the current 2018 UNOS Adult Heart Allocation Policy, we assessed whether there are significant regional differences in the rates of sex-mismatching across UNOS regions using the proportion test and further assessed whether the rates of utilization of sex-mismatching in males is correlated with the rates of sex-mismatching in females across regions.

Post-transplant survival was analyzed using the Kaplan-Meier method and was compared between groups via the Wilcoxson log-rank test. We also estimated Restricted Mean Survival Time (RMST) and between-arm survival differences in RMST.

For binary outcomes, we performed logistic regression to examine the effects of sex-mismatch on males and females for the postoperative outcomes of pre-discharge stroke, pre-discharge dialysis, and treatment for acute rejection within one year.

All statistical computing was completed using R version 4.3.0 (2023-04-21) (The R Foundation, Vienna, Austria).^10^ We performed propensity score matching using MatchIt version 4.5.3. We estimated RMST using survRM2 version 1.0.4.

### Equivalence Testing

For binary outcomes, our primary measure of effect size is the odds ratio. We selected an a priori equivalence margin in line with Wellek’s strict equivalence margin on the log-odds scale of 0.41, which corresponds to testing whether the odds ratio between arms is contained within the interval (0.66—1.51).^11^ The choice of this equivalence margin is in line with the discussion of what a large odds ratios discussed in Chen et al.^12^ This equivalence margin was chosen prior to performing statistical analysis.

For survival outcomes, our primary measure of between-arm effect size was the between-arm RMST ratio. We considered a margin on the log-ratio scale of 0.05, which corresponds to a between-arm RMST ratio within (0.95—1.05), to be a clinically relevant difference. These bounds correspond roughly to a tolerance of between-arm differences of days at 30 days, ±18 days at one year, ±91 days at five years, and ±182 days at ten years. This equivalence margin was chosen prior to performing statistical analysis.

We reported the p-values corresponding to two-sided significant differences and Two One-Sided Testing (TOST) equivalence, denoted P_Difference_ and P_Equivalence_ respectively. All confidence intervals two-sided Wald confidence intervals.

### Sensitivity Analysis: The Use of Predicted Heart Mass

PHM is a commonly used estimate of heart size in heart transplantation. It was developed in 2019 as a model of adult heart size to facilitate donor-recipient size matching.^8^ A patient’s PHM is a function of age, weight, height, and sex. This measure refines other notions of donor-recipient size mismatch such as body mass index, body surface area, and weight mismatch. A donor-recipient ratio of <0.86 is considered undersized. ^8^ It has been widely reported that undersizing of the heart confers risk to heart transplant recipients.^6,8,13,14,15,16,17^ To ensure that our findings were robust with respect to other notions of size-mismatching, we performed three separate matched analyses with respect to donor-recipient BMI ratio, body surface area ratio, and weight ratio (Figures S6, S7, and S8).

### IRB Exemption

This study is a retrospective review of deidentified data supplied by UNOS in the form of a deidentified Standard Research Analysis File. Our project was determined to be Institutional Review Board exempt following a human subjects research determination (protocol #71189).

## Results

### Baseline Characteristics

A total of 53,595 male isolated heart transplant recipients met inclusion criteria. The mean age was 53.3 years (SD: 11.6). 11,149 (20.8%) male recipients were sex-mismatched and 42,446 (79.2%) were sex-matched. The donor-recipient PHM ratios of the male sex-matched and the sex-mismatched patients were 1.02 and 0.85, respectively. A total of 17,015 female isolated heart transplant recipients met inclusion criteria. The mean age was 49.6 years (SD: 13.0). 7,572 (44.5%) female recipients were sex-mismatched and 9,443 (55.5%) were sex-matched. The donor-recipient PHM ratios of the female sex-matched cohort and the sex-mismatched were 1.06 and 1.31, respectively. Comprehensive baseline characteristics for the unmatched cohorts are shown in Table 1.

After propensity score matching, 3,768 pairs of male recipients with and without donor-recipient sex-mismatch were identified. The donor-recipient PHM ratios of the sex-matched and the sex-mismatched male recipients were 0.93 and 0.91, respectively. Additionally, 2,266 pairs of female recipients with and without donor-recipient sex-matched were identified. The donor-recipient PHM ratios of the sex-matched and the sex-mismatched patients were 1.19 and 1.23, respectively. The propensity matched variables were well balanced in both SMD (all below 0.125 for male and 0.19 for female recipients) and variance ratio (between 0.82 and 1.16 for male recipients and 0.98 and 1.35 for female recipients).

### Utilization of Sex-Mismatched Heart Allografts Across Time and Geography

Over the study period, the utilization of sex-mismatched donor allograft decreased over time (male Spearman correlation coefficient *ρ*=-0.78, p=3.8e-07; female Spearman correlation coefficient *ρ*=-0.87, p<2e-12, Figure 1). There was also significant regional variation in the utilization of sex-mismatched donor allograft (p<0.001 for both male and female recipients, Figure 2) among those transplanted after the 2018 UNOS Adult Heart Allocation Policy revisions.

**Figure 1:**
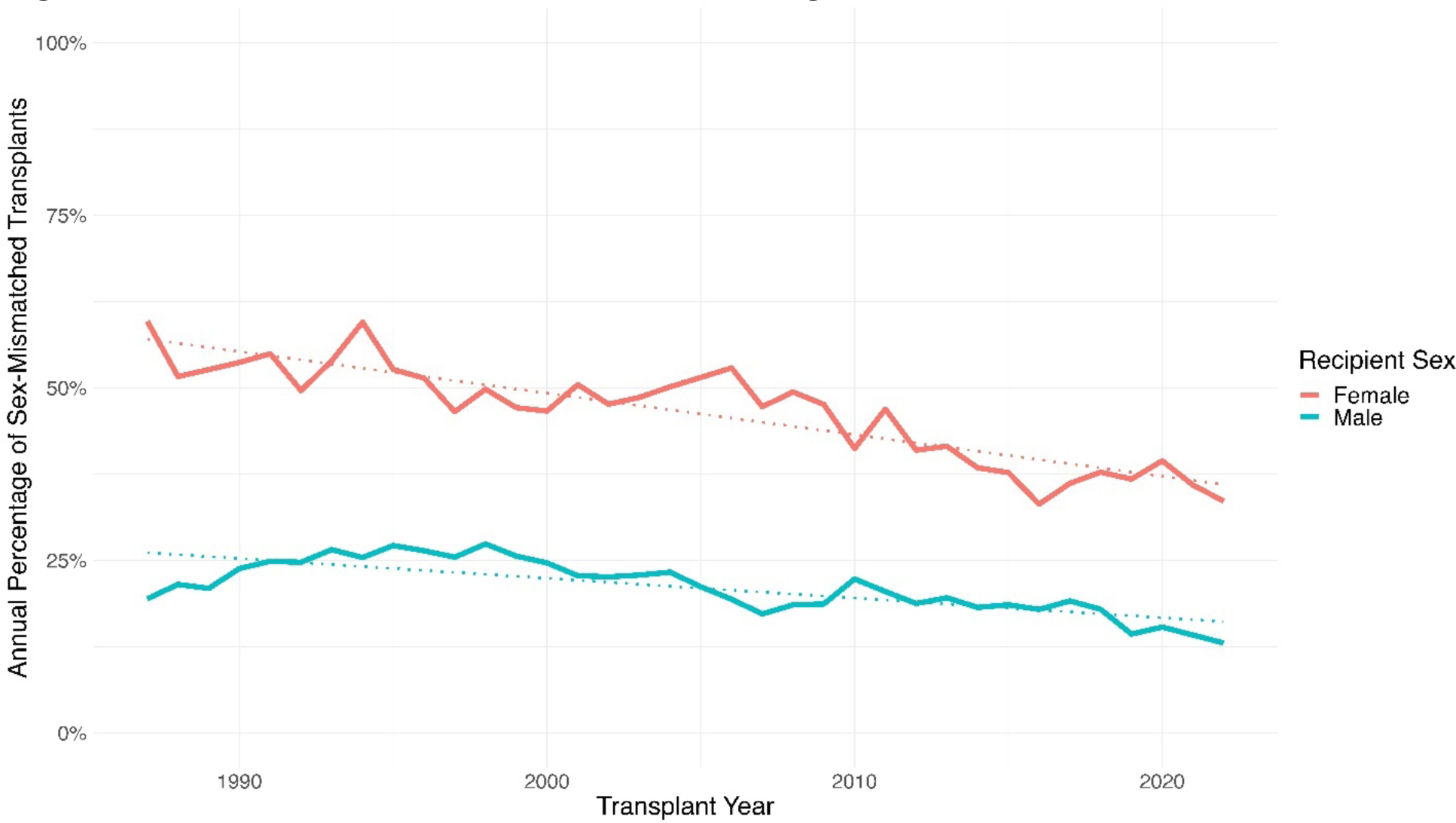
Trends in Utilization of Sex-Mismatching.

**Figure 2:**
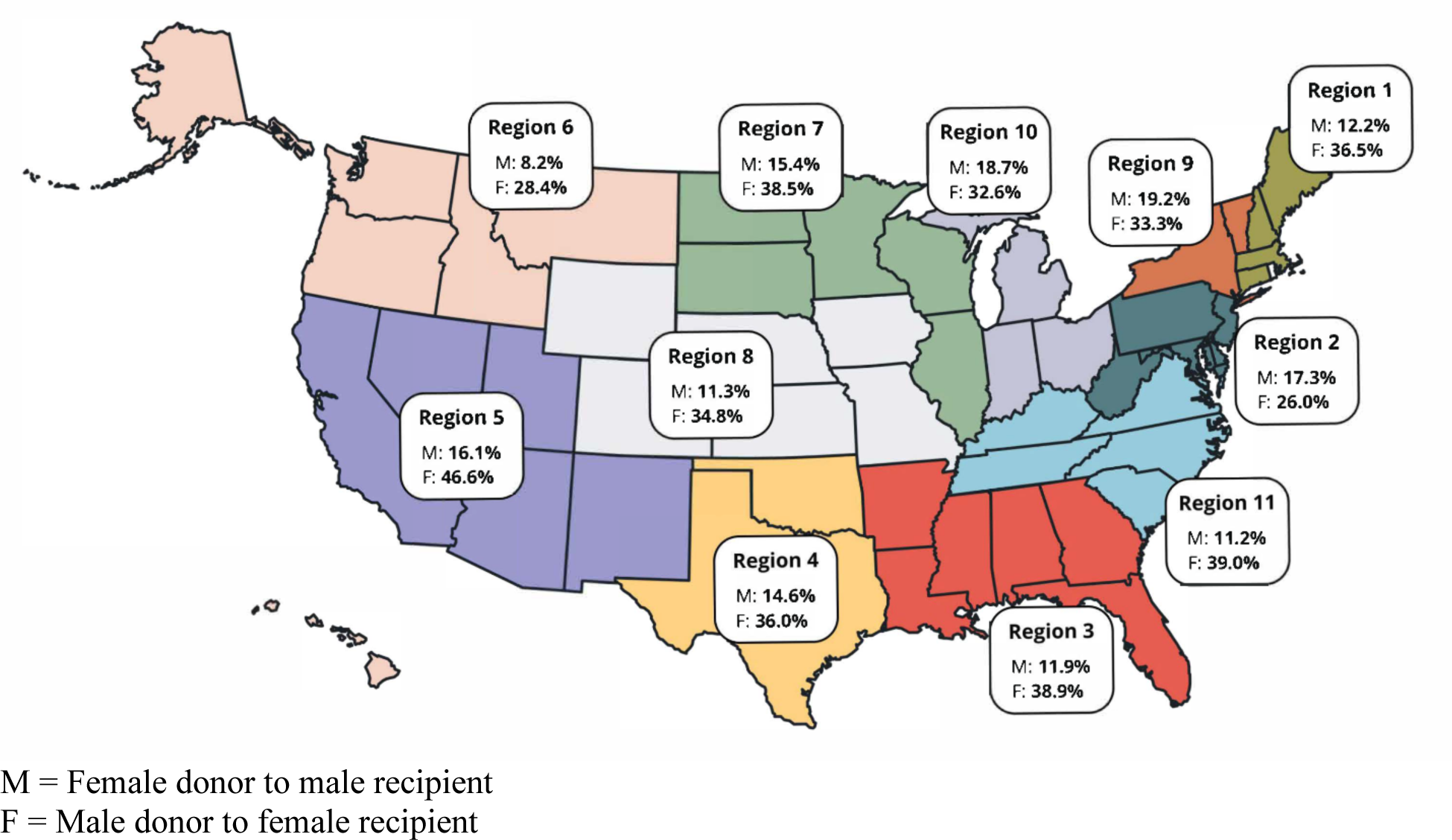
Percentage of Sex-Mismatched Isolated Heart Transplants Since 2018 by UNOS Region.

Since the UNOS Heart Allocation Policy change in 2018, we observed significant differences between the proportion of sex-mismatched allografts given to male recipients across UNOS regions (P<0.001) and to female recipients (P<0.001) as visualized in Figure 2. A striking example of this is seen in the difference between UNOS Region 5 and UNOS Region 6. Region 5 had a higher rate of sex-mismatched allografts for both male and female recipients compared to the neighboring UNOS Region 6 (16.1% versus 8.2% for male recipients and 46.6% versus 28.4% for female recipients, respectively). Further information is reported in the supplement (Table S8 for male recipients, Table S9 for female recipients).

To assess whether there was a regional cross-correlation between the respective rates of sex-mismatching for male and female recipients, we ranked UNOS regions in two ways: first in descending order of the proportion of sex-mismatched males recipients and second in descending order of the proportion of sex-mismatched female recipients. We found a small and statistically insignificant correlation between these rankings (Spearman correlation coefficient *ρ*=-0.19, P=0.57). It appears that the rate of mismatching for males is not correlated with the sex-mismatching for females within each region.

### Analysis of Post-transplant Outcomes

In the propensity-matched cohort, there was no significant difference in overall survival between sex-matched and sex-mismatched groups for male recipients (P=0.71) or female recipients (P=0.27, Figure 3). We inferred significant equivalence in RMST between sex-matching strata for both male and female recipients at all temporal endpoints from 30 days to 10 years (P_equivalence_<0.003 for all temporal endpoints).

**Figure 3:**
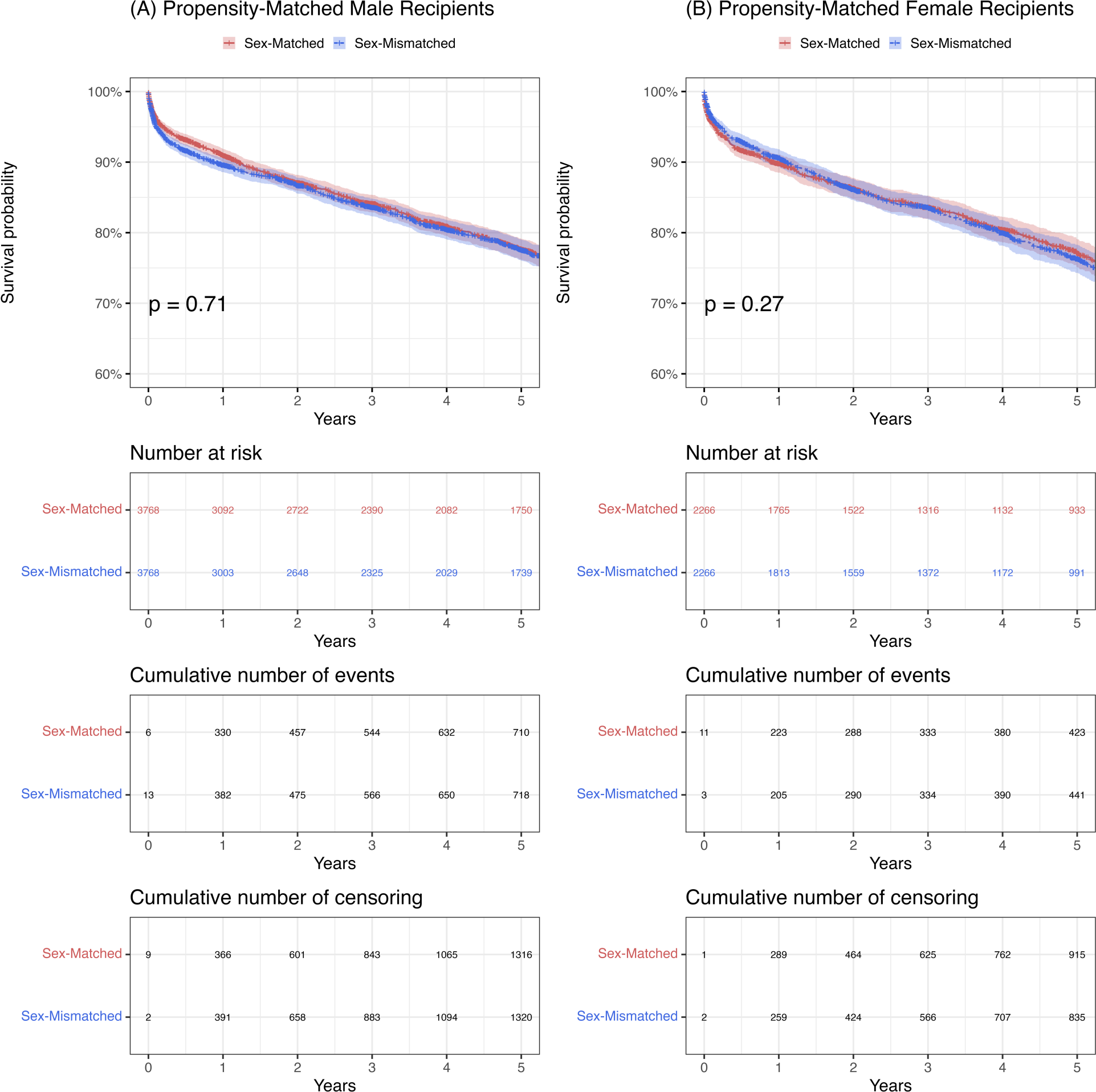
Kaplan-Meier Analysis of Propensity-Matched Heart Transplant Recipients Stratified by Sex-matching Status.

**Figure 4:**
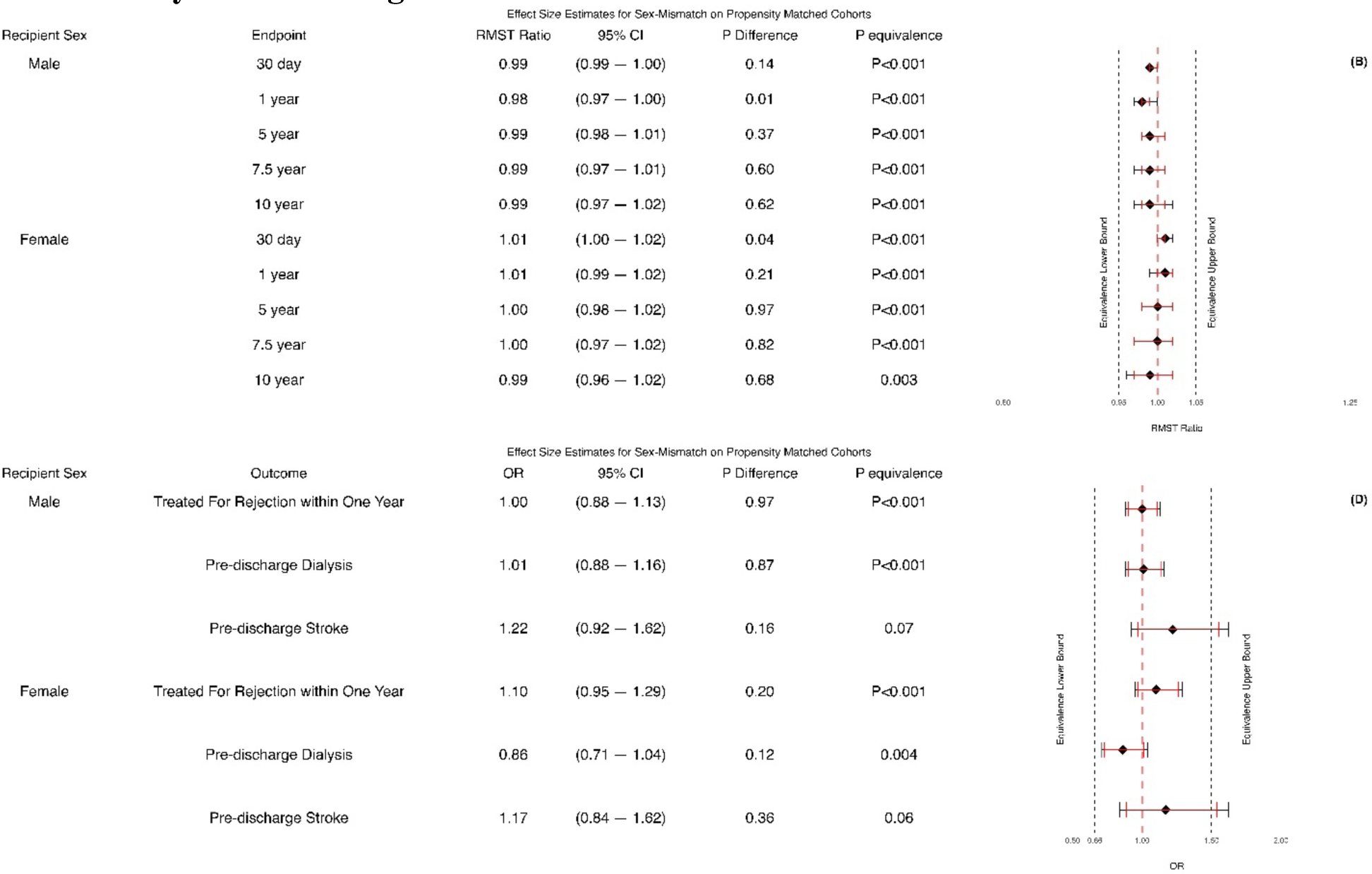
Postoperative Outcomes for Propensity-Matched Heart Transplant Recipients Stratified by Sex-matching Status.

For secondary outcomes, we inferred significant equivalence in treatment for rejection within one year (P_equivalence_<0.001 for both male and female) and pre-discharge dialysis (male: P_equivalence_<0.001; female: P_equivalence_=0.004). Neither significant equivalence nor significant difference were inferred for pre-discharge stroke (male: P_equivalence_=0.06; female: P_equivalence_=0.07). Further details can be found in Table S7.

## Discussion

In this analysis of a national heart transplant registry, we observed declining rates of sex-mismatching for both male and female recipients since 1987. In the current era, since the 2018 UNOS Adult Heart Allocation Policy change, we observed large differences across regions in the utilization of sex-mismatched allografts. More importantly, donor-recipient sex-mismatch confers no marginal risk to the adverse postoperative outcomes when other donor and recipient risk factors were controlled, including donor and recipient size matching. Given these findings, sex-mismatching should not be considered an independent cause for concern when determining whether to accept or reject a donor heart.

However, this does not mean that sex-mismatched allografts are risk-free. Due to base rate differences in heart sizing between sexes, the probability of a randomly selected sex-mismatched donor-recipient pair being compatible is far less than the probability of a randomly selected sex-matched pair being compatible. We acknowledge that severe undersizing of heart allografts poses a risk to patients, but our analysis has shown that a size-compatible sex-mismatched allograft would be equivalent in clinical outcomes to a sex-matched allograft of otherwise identical covariates.

This result suggests a strategy to improve access to heart transplant by expanding the donor pool. Female donors are potentially underutilized and represent less than 30% of donor pool since the 2018 UNOS Adult Heart Allocation Policy change. Lo et al. suggest that removing phenotypic constraints to matching improves average waitlist time by expanding the donor pool.^18^ Even though males make up the majority of heart transplant recipients, most male recipients are compatible with over a quarter of female donors (Table S2). Unfortunately, our analysis was limited to considering only donors who were ultimately transplanted, and further research is needed to quantify how many more female donors can be added to the donor pool altogether.

### The Role of Compatibility and Phenotypic Matching in Organ Allocation

A distinctive feature of transplantation is the need to assess the compatibility of a given donor allograft with the recipient. It has recently been argued that there is no such thing as the “perfect donor,” and that adding undue phenotypic donor-recipient compatibility constraints negatively impacts the prospects of waitlisted patients by restricting the donor pool of “compatible” donors.^5^ Therefore, it is imperative to identify the optimal compatibility constraints to balance the likelihood of receiving an organ while mitigating the risk of adverse postoperative outcomes due to receiving an incompatible organ.

Sex-mismatching has been proposed as a risk factor for adverse postoperative outcomes with various causal mechanisms proposed.^19,20^ However, our results suggest that all of the associated risk is accounted for by other known risk factors.

### Limitations

The primary limitation of this study is that the UNOS database does not record or provide actual measurements of heart size. Therefore we are limited in using PHM, an estimate of heart mass. It has been argued that PHM is the most useful estimate of heart size.^8^ Further work on sufficiently large cohorts with direct measurements of heart size would provide further insight into the role that heart sizing and adjustment on known risk factors plays in postoperative outcomes.

Second, in the all-comers cohort, the causal positivity assumption was violated due to the strong association between donor-recipient PHM ratio and sex-mismatching for both male and female recipients. As a result, sex-mismatched males with very low donor-recipient PHM ratio and sex-mismatched females with very high donor-recipient PHM ratio were unmatched. This is an example of what Zhu et al. term a “practical violation” of the causal positivity assumption.^21^

Finally, there is always the potential that the propensity model used in matching is misspecified, either by a suboptimal choice of variables or through the choice of model used to estimate the propensity score.^22^

### Conclusion

Our analysis revealed significant equivalence between sex-matched and sex-mismatched recipients of isolated heart transplant on primary outcomes of survival up to 10 years and secondary outcomes of one-year acute rejection and pre-discharge dialysis in both female and male recipients when properly matched for heart size. We therefore recommend that sex-mismatch not be factored into heart donor acceptance decisions. Moreover, sex-mismatching should not be a factor considered in evaluating the compatibility of a recipient in an updated organ allocation policy. We hope that the female donor acceptance rate can be increased to allow higher utilization if there is an appropriately heart size matched recipient available.

## Data Availability

Dr. Reid Dale had full access to all the data in the study and takes responsibility for its integrity and the data analysis.

## Glossary of Abbreviations

OR: Odds Ratio
UNOS: United Network for Organ Sharing
PHM: Predicted Heart Mass
BMI: Body Mass Index
SMD: Standardized Mean Difference
RMST: Restricted Mean Survival Time
TOST: Two One-Sided Test
SD: Standard Deviation

## Notes

### Competing Interest Statement

The authors have declared no competing interest.

### Funding Statement

The authors have no funding sources to disclose.

